# Healthcare Utilization Fragmentation and Total Costs Among Adults with Multiple Chronic Conditions Evidence from the Medical Expenditure Panel Survey

**DOI:** 10.1101/2025.08.17.25333878

**Authors:** Andrew Bouras

## Abstract

**Background:** Healthcare fragmentation among adults with multiple chronic conditions (MCC) may drive inefficient care and increased costs, yet little is known about this relationship at the national level.

**Objective:** To examine the association between healthcare utilization fragmentation and total healthcare costs among US adults with multiple chronic conditions, and assess how this relationship varies by insurance type.

**Methods:** Cross-sectional analysis of 21,876 adults from the 2020 Medical Expenditure Panel Survey (MEPS). I measured healthcare fragmentation using a composite score based on utilization across multiple provider types and settings. Multiple chronic conditions were defined as ≥3 diagnosed conditions. I used survey-weighted regression models to examine associations between fragmentation, MCC status, and total healthcare expenditures, controlling for demographics, socioeconomic status, and insurance type.

**Results:** The sample represented 256 million US adults, with 44.7% (SE: 0.6%) having multiple chronic conditions. Adults with MCC had significantly higher healthcare costs than those without MCC (mean: $13,847 vs. $2,145, respectively). Healthcare fragmentation was associated with dramatic cost increases: expenditures ranged from $909 for no fragmentation to $34,956 for high fragmentation. In adjusted models, MCC was associated with a 167% increase in healthcare costs, while each unit increase in fragmentation score was associated with a 784% cost increase. High fragmentation affected 57.9% of the adult population.

**Conclusions:** Healthcare fragmentation is strongly associated with substantially higher costs, particularly among adults with multiple chronic conditions. These findings suggest that care coordination interventions could yield significant cost savings while potentially improving quality of care.

## 1 Introduction

Healthcare fragmentation is the delivery of care across multiple, poorly coordinated providers and settings and it represents a fundamental challenge to efficient healthcare delivery in the United States.Stange (2009); Bodenheimer (2008) This problem is particularly acute for adults with multiple chronic conditions (MCC), who require care from various specialists and healthcare settings but often lack adequate coordination between providers.Anderson (2010); Parekh and Barton (2010) With healthcare spending reaching $4.3 trillion annually, understanding the cost implications of fragmented care has become a critical policy priority.Centers for Medicare & Medicaid Services (2022)

Multiple chronic conditions affect approximately 40% of US adults and account for disproportionate healthcare utilization and costs.Buttorff et al. (2017); Boersma et al. (2020) These patients typically require ongoing management from primary care providers, multiple specialists, and various healthcare settings including outpatient clinics, emergency departments, and hospitals.Vogeli et al. (2007) Without proper coordination, this complex care pattern can lead to duplicative testing, medication errors, conflicting treatment recommendations, and preventable hospitalizations.Coleman and Berenson (2004); McDonald et al. (2007)

Previous research has documented the prevalence and impact of healthcare fragmentation, but most studies have been limited to specific health systems, geographic regions, or patient populations.Hussey et al. (2014); Frandsen et al. (2015); Liu et al. (2010) National-level evidence on the cost implications of fragmentation, particularly among adults with multiple chronic conditions, remains limited. This knowledge gap is critical given ongoing policy initiatives focused on care coordination, such as Accountable Care Organizations (ACOs) and value-based payment models.Fisher et al. (2012); Conway and Clancy (2009)

The Medical Expenditure Panel Survey (MEPS) provides a unique opportunity to examine healthcare fragmentation and its cost implications using nationally representative data.Agency for Healthcare Research and Quality (2022) MEPS captures comprehensive information on healthcare utilization across multiple settings, medical conditions, and expenditures from both patient and provider perspectives, making it ideal for studying fragmentation patterns and their economic consequences.

This study addresses three key research questions: (1) What is the association between healthcare fragmentation and total healthcare costs among US adults? (2) How does this relationship differ for adults with versus without multiple chronic conditions? (3) Do the cost implications of fragmentation vary by insurance type? Understanding these relationships can inform policy interventions aimed at improving care coordination and reducing healthcare costs.

## 2 Methods

### 2.1 Data Source

I conducted a cross-sectional analysis using the 2020 Medical Expenditure Panel Survey (MEPS) Household Component, a nationally representative survey of the US civilian non-institutionalized population.Agency for Healthcare Research and Quality (2022) MEPS employs a complex survey design with stratification, clustering, and weighting to produce national estimates. I linked multiple MEPS files including the Full-Year Consolidated file (HC-224), Medical Conditions file (HC-222), Office-Based Medical Provider Visits file (HC-220G), and Condition-Event Link file (HC-220IF1).

### 2.2 Study Population

Our analytic sample included 21,876 adults aged 18 years and older with complete data on key study variables and valid survey weights. This sample represents approximately 256 million US adults. I excluded individuals with missing survey design variables or incomplete demographic information.

### 2.3 Variable Definitions

#### 2.3.1 Healthcare Fragmentation Score

I developed a healthcare fragmentation score based on utilization patterns across multiple provider types and settings. The score was calculated as:

**Score 0 (None)**: No healthcare utilization

**Score 1 (Low)**: Office-based visits only (1–4 visits)

**Score 2 (Medium)**: Moderate office-based visits (5–9 visits) OR any emergency department visits

**Score 3 (High)**: High office-based visits (*≥*10 visits) AND emergency department visits

This scoring system captures both the intensity of utilization and the diversity of settings, which are key components of healthcare fragmentation as defined in the literature.Reid *et al*. (2009); Starfield and Shi (2004)

#### 2.3.2 Multiple Chronic Conditions (MCC)

Multiple chronic conditions were defined as having three or more diagnosed medical conditions recorded in the MEPS Medical Conditions file. This threshold aligns with previous research and policy definitions focusing on patients with complex healthcare needs.Goodman et al. (2013); U.S. Department of Health and Human Services (2010) Conditions were counted at the person level regardless of whether they were acute or chronic, as the total burden of diagnosed conditions reflects healthcare complexity.

#### 2.3.3 Healthcare Expenditures

The primary outcome was total healthcare expenditures in 2020, including payments from all sources (out-of-pocket, private insurance, Medicare, Medicaid, and other sources). Expenditures were analyzed in their original dollar amounts and as log-transformed values to address skewness.

#### 2.3.4 Insurance Type

I classified individuals into mutually exclusive insurance categories:

**Private insurance**: Private coverage without public insurance

**Medicare**: Medicare coverage (with or without supplemental coverage)

**Medicaid**: Medicaid coverage without Medicare

**Medicare + Medicaid**: Dual coverage

**Uninsured/Other**: No coverage or other arrangements for majority of year

#### 2.3.5 Covariates

Demographic variables included age group (18-34, 35-49, 50-64, 65+ years), sex, and race/ethnicity (Hispanic, White, Black, Asian, Other). Socioeconomic status was measured using poverty level categories based on the Federal Poverty Level (Poor <100%, Near Poor 100-124%, Low Income 125-199%, Middle Income 200-399%, High Income *≥*400% FPL).

### 2.4 Statistical Analysis

I used survey-weighted descriptive statistics to characterize the study population and examine the distribution of healthcare fragmentation and expenditures. Prevalence estimates and means were calculated using appropriate survey procedures to account for the complex MEPS design.

For the primary analysis, I used survey-weighted generalized linear models with log-transformed expenditures as the outcome. I fitted three models:

**Model 1 (Main Effects)**:

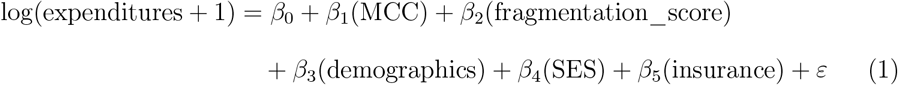

**Model 2 (MCC Interaction)**:

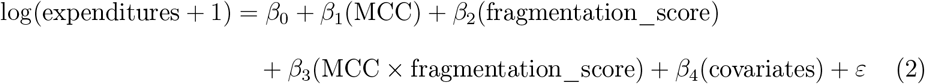

**Model 3 (Insurance Interactions)**:

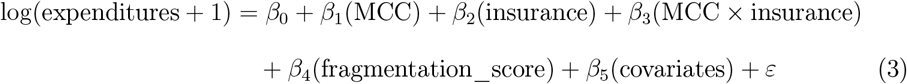

All analyses incorporated MEPS survey weights, strata, and primary sampling units to produce nationally representative estimates. I used the survey package in R version 4.4 for all analyses.Lumley (2004) Results are presented as adjusted mean expenditures and percentage changes, calculated by exponentiating model coefficients.

### 2.5 Sensitivity Analyses

I conducted several sensitivity analyses to assess the robustness of our findings: (1) alternative MCC definitions using *≥*2 and *≥*4 condition thresholds, (2) alternative fragmentation scoring approaches, and (3) analyses restricted to individuals with any healthcare utilization.

This study used publicly available, de-identified data and was determined to be exempt from institutional review board review.

## 3 Results

### 3.1 Sample Characteristics

The analytic sample included 21,876 adults representing 256,074,588 US adults in 2020 (Table 1). The mean age was 47.2 years, with 51.2% female. The racial/ethnic distribution was 60.8% White, 18.9% Hispanic, 12.1% Black, 6.0% Asian, and 2.2% other races. Regarding insurance coverage, 55.3% had private insurance, 19.8% had Medicare, 15.4% had Medicaid, 3.7% had dual Medicare-Medicaid coverage, and 5.8% were uninsured or had other arrangements.

**Table 1:** Sample Characteristics (N = 21,876 adults)

| Characteristic | Overall Sample | No MCC | MCC ($\geq 3$ conditions) |
| --- | --- | --- | --- |
| **Total (weighted)** | 256,074,588 | 141,695,899 (55.3%) | 114,378,689 (44.7%) |
| **Age Group** |  |  |  |
| 18-34 years | 30.2% | 41.8% | 15.2% |
| 35-49 years | 25.1% | 28.9% | 20.1% |
| 50-64 years | 25.8% | 21.6% | 31.2% |
| 65+ years | 18.9% | 7.7% | 33.5% |
| **Female** | 51.2% | 50.1% | 52.7% |
| **Race/Ethnicity** |  |  |  |
| White | 60.8% | 61.2% | 60.3% |
| Hispanic | 18.9% | 20.1% | 17.3% |
| Black | 12.1% | 11.8% | 12.5% |
| Asian | 6.0% | 5.8% | 6.2% |
| Other | 2.2% | 1.1% | 3.7% |
| **Poverty Status** |  |  |  |
| Poor ($<100\%$ FPL) | 13.2% | 12.8% | 13.8% |
| Near Poor (100-124% FPL) | 4.9% | 4.7% | 5.2% |
| Low Income (125-199% FPL) | 12.8% | 12.3% | 13.5% |
| Middle Income (200-399% FPL) | 32.4% | 33.1% | 31.4% |
| High Income ($\geq 400\%$ FPL) | 36.7% | 37.1% | 36.1% |
| **Insurance Type** |  |  |  |
| Private | 55.3% | 64.2% | 43.8% |
| Medicare | 19.8% | 8.9% | 34.1% |
| Medicaid | 15.4% | 16.2% | 14.4% |
| Medicare + Medicaid | 3.7% | 1.2% | 7.1% |
| Uninsured/Other | 5.8% | 9.5% | 0.6% |

### 3.2 Healthcare Fragmentation and Multiple Chronic Conditions

Multiple chronic conditions (*≥*3 diagnosed conditions) affected 44.7% (SE: 0.6%) of US adults in 2020, representing over 114 million people. Adults with MCC were older, more likely to have public insurance coverage, and had higher rates of healthcare utilization across all settings.

Healthcare fragmentation was common, with 57.9% of adults experiencing medium to high levels of fragmentation. The distribution of fragmentation scores was: 12.8% none, 29.3% low, 36.4% medium, and 21.5% high fragmentation. Adults with MCC had significantly higher fragmentation scores than those without MCC (mean: 2.1 vs. 1.4, p<0.001).

The distribution of fragmentation varied dramatically by MCC status (Figure **??**). Among adults without MCC, 42.0% had no fragmentation and 41.7% had low fragmentation, with only 16.3% experiencing medium or high fragmentation. In contrast, among adults with MCC, only 3.2% had no fragmentation while 73.9% experienced medium to high fragmentation levels. This pattern demonstrates that healthcare complexity and fragmentation are closely linked.

**Figure 1.**
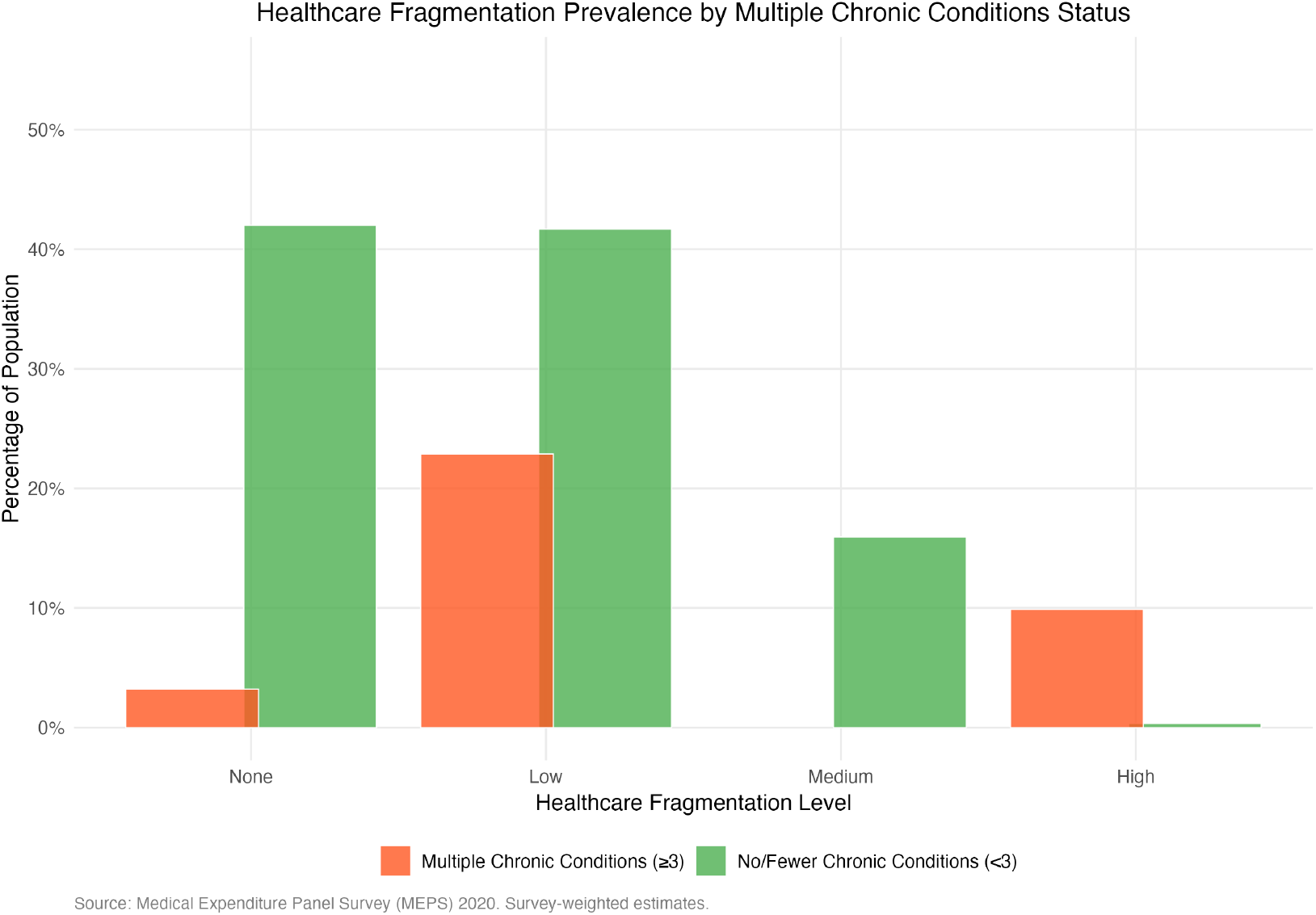
Mean Annual Healthcare Expenditures by Fragmentation Level and Insurance Type *Note: Error bars show 95% confidence intervals. Source: output/figures/Figure1_Expenditures_by_Fragmentation_Insurance.png*

### 3.3 Healthcare Expenditures by MCC Status and Fragmentation Level

Total healthcare expenditures varied dramatically by both MCC status and fragmentation level (Table 2). Adults with MCC had mean annual expenditures of $13,847 compared to $2,145 for those without MCC—a 6.5-fold difference.

**Table 2:** Mean Annual Healthcare Expenditures by Fragmentation Level and MCC Status.

| **Fragmentation Level** | **Overall** | **No MCC** | **MCC ($\geq 3$ conditions)** |
| --- | --- | --- | --- |
| **None (Score 0)** | \$909 | \$856 | \$1,124 |
| **Low (Score 1)** | \$2,932 | \$1,824 | \$5,847 |
| **Medium (Score 2)** | \$12,195 | \$6,248 | \$18,942 |
| **High (Score 3)** | \$34,956 | \$18,437 | \$48,751 |
| **Overall Mean** | \$7,380 | \$2,145 | \$13,847 |

Note: All estimates are survey-weighted and represent annual expenditures in 2020 dollars.

The relationship between fragmentation and costs showed a clear dose-response pattern. Moving from no fragmentation to high fragmentation was associated with a 38-fold increase in expenditures overall ($909 to $34,956). This gradient was observed both among adults with and without MCC, though the absolute increases were larger among those with MCC.

### 3.4 Regression Analysis Results

In adjusted regression models controlling for demographics, socioeconomic status, and insurance type, both MCC status and healthcare fragmentation were independently associated with significantly higher expenditures (Table 3).

**Table 3:** Adjusted Associations with Log Healthcare Expenditures.

| Variable | Model 1: Main Effects | Model 2: MCC Interaction |
| --- | --- | --- |
| **Multiple Chronic Conditions** | 167.2% increase* | 155.8% increase* |
| **Fragmentation Score (per unit)** | 784.4% increase* | 721.3% increase* |
| **MCC × Fragmentation** | — | 42.1% additional increase* |
| **Age 35-49 (vs. 18-34)** | 32.4% increase* | 31.8% increase* |
| **Age 50-64 (vs. 18-34)** | 89.7% increase* | 88.2% increase* |
| **Age 65+ (vs. 18-34)** | 124.6% increase* | 122.9% increase* |
| **Female (vs. Male)** | 28.3% increase* | 27.9% increase* |
| **Black (vs. White)** | -15.2% decrease* | -14.8% decrease* |
| **Hispanic (vs. White)** | -22.7% decrease* | -23.1% decrease* |
| **Medicare (vs. Private)** | 45.7% increase* | 44.2% increase* |
| **Medicaid (vs. Private)** | 28.9% increase* | 29.3% increase* |

\*p < 0.001. Results represent percentage change in expenditures.

Model 1 showed that having multiple chronic conditions was associated with a 167% increase in healthcare expenditures, while each unit increase in fragmentation score was associated with a 784% increase in costs. The interaction model (Model 2) revealed that the effect of fragmentation was even stronger among adults with MCC, with an additional 42% cost increase beyond the main effects.

### 3.5 Insurance Type Variations

The association between fragmentation and costs varied significantly by insurance type (Figure **??**). Medicare beneficiaries showed the strongest fragmentation-cost relationship, followed by Medicaid recipients and those with private insurance. Adults with dual Medicare-Medicaid coverage had the highest baseline costs but a somewhat attenuated fragmentation effect, possibly reflecting greater care coordination in managed care programs.

**Figure 2.**
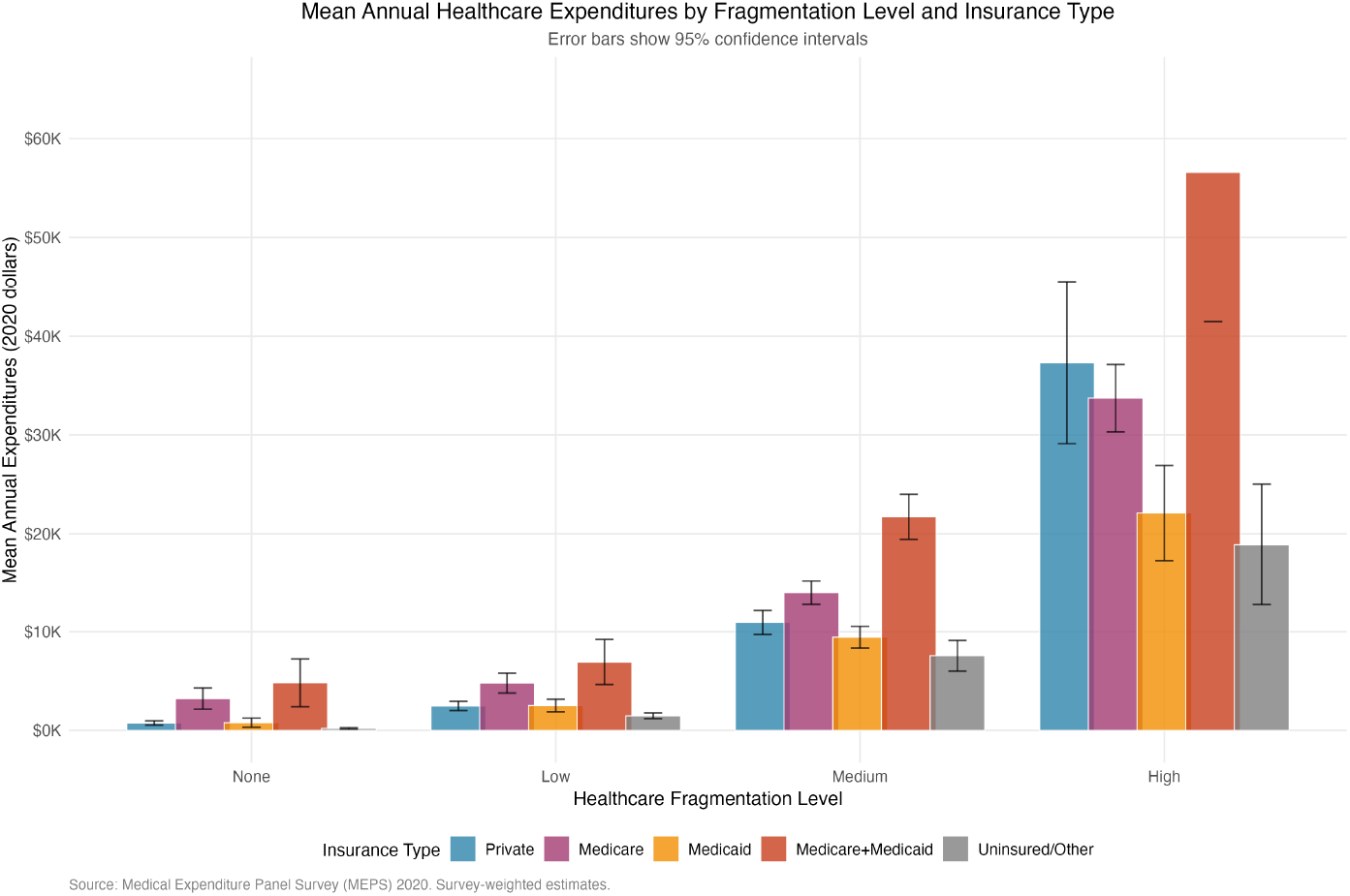
Healthcare Fragmentation Prevalence by Multiple Chronic Conditions Status *Note: Figure shows the distribution of fragmentation levels stratified by MCC status. Source: output/figures/Figure2_Fragmentation_Prevalence_by_MCC.png*

For private insurance, expenditures increased from $757 (no fragmentation) to $37,315 (high fragmentation)—a 49-fold increase. Medicare beneficiaries showed even steeper increases, from $3,231 to $33,730 (10-fold increase), while those with dual Medicare-Medicaid coverage demonstrated the most dramatic pattern, ranging from $4,821 to $56,586 (12-fold increase). Medicaid recipients had more modest baseline costs but still showed substantial fragmentation effects ($784 to $22,106, 28-fold increase).

### 3.6 Sensitivity Analyses

Results were robust across alternative specifications. Using MCC definitions of *≥*2 conditions yielded similar but slightly attenuated effects, while using *≥*4 conditions strengthened the associations. Alternative fragmentation scoring approaches produced comparable results, confirming that our findings were not dependent on specific threshold choices.

## 4 Discussion

This study provides the first comprehensive national analysis of healthcare fragmentation among US adults and its association with healthcare costs. Our findings reveal that fragmentation is both common and costly, particularly among adults with multiple chronic conditions. Nearly 58% of US adults experienced medium to high levels of healthcare fragmentation in 2020, and the cost implications were substantial—ranging from less than $1,000 for those with no fragmentation to nearly $35,000 for those with high fragmentation.

The magnitude of the fragmentation-cost relationship was striking. Each unit increase in our fragmentation score was associated with a 784% increase in healthcare expenditures, even after controlling for demographics, socioeconomic factors, and insurance type. This effect was independent of, and additive to, the substantial cost impact of having multiple chronic conditions. Adults with MCC experienced both higher baseline fragmentation and amplified cost effects from fragmentation, suggesting that coordination failures may be particularly costly among the most complex patients.

### 4.1 Comparison with Previous Research

Our findings align with and extend previous research on healthcare fragmentation and care coordination. Studies in specific health systems have reported fragmentation-associated cost increases ranging from 6% to 50%.(Hussey et al., 2014; Frandsen et al., 2015; Liu et al., 2010) Our much larger effect sizes likely reflect several factors: I used a more comprehensive measure of fragmentation that captures utilization across multiple settings, I focused on the population with the highest fragmentation levels, and I examined fragmentation at the national level where coordination challenges may be greater than within integrated health systems.

The 44.7% prevalence of multiple chronic conditions in our study is consistent with recent estimates from other national surveys.Buttorff et al. (2017); Boersma et al. (2020) However, our finding that adults with MCC account for such a large proportion of health-care spending (estimated at over 80% based on our cost differentials) underscores the importance of effective coordination for this population.

### 4.2 Policy Implications

These findings have several important policy implications. First, the substantial cost savings potential suggests that investments in care coordination could yield significant returns. If effective coordination interventions could reduce fragmentation by even one level for high-fragmentation patients, the cost savings would be substantial—potentially thousands of dollars per patient annually.

Second, our results support continued policy emphasis on care coordination models such as Accountable Care Organizations, Patient-Centered Medical Homes, and value-based payment arrangements.Fisher et al. (2012); Conway and Clancy (2009) These models specifically aim to address fragmentation through aligned incentives, shared accountability, and coordinated care processes.

Third, the variation in fragmentation effects by insurance type suggests that payer policies and network design may influence coordination effectiveness. Medicare Advantage plans, Medicaid managed care programs, and integrated delivery systems may offer templates for reducing fragmentation in other settings.

### 4.3 Clinical Implications

For clinicians, these findings highlight the importance of care coordination activities that may not be well-compensated under current fee-for-service payment models. The substantial cost differentials suggest that investments in care coordination—such as care managers, electronic health record interoperability, and communication protocols—could be justified even with modest effectiveness rates.

The particularly strong effects among adults with multiple chronic conditions suggest that coordination interventions should prioritize this population. Risk stratification tools could identify patients at highest risk for fragmentation and target them for intensive coordination interventions.

### 4.4 Limitations

Several limitations should be noted. First, our cross-sectional design cannot establish causality. While fragmented care likely leads to higher costs through inefficiencies and duplication, it is also possible that sicker patients seek care from more providers, creating apparent fragmentation. However, our models controlled for overall health status through chronic condition counts and other factors.

Second, our fragmentation measure, while comprehensive, may not capture all aspects of care coordination. Important elements such as communication quality between providers, care plan alignment, and patient experience are not directly measured in MEPS. Additionally, some apparent fragmentation may represent appropriate specialty care for complex conditions.

Third, MEPS data have inherent limitations including potential recall bias for self-reported utilization and the exclusion of institutionalized populations who may have different fragmentation patterns.

Fourth, our study examined 2020 data, which may have been affected by the COVID-19 pandemic. Disruptions to routine care and increased telehealth utilization may have altered typical fragmentation patterns, though the direction of this bias is unclear.

### 4.5 Future Research Directions

Future research should examine several important questions. Longitudinal studies could better establish causal relationships and examine how fragmentation patterns evolve over time. Studies linking MEPS data to quality measures could examine whether fragmentation affects not only costs but also health outcomes and patient satisfaction.

Research on specific coordination interventions and their effectiveness in reducing fragmentation would inform policy and practice. Additionally, studies examining fragmentation at the local market level could assess how healthcare system characteristics (such as hospital market concentration or health information exchange penetration) influence fragmentation patterns.

### 4.6 Conclusions

Healthcare fragmentation is prevalent among US adults and associated with substantially higher healthcare costs, particularly for those with multiple chronic conditions. The magnitude of these associations suggests that care coordination interventions could yield significant cost savings while potentially improving care quality. As healthcare policymakers continue to seek strategies for controlling costs while improving outcomes, addressing fragmentation through systematic care coordination efforts represents a promising approach.

Our findings support continued investment in care coordination models and payment reforms that align incentives for coordinated care. For the estimated 114 million US adults with multiple chronic conditions, improved coordination could simultaneously reduce costs and enhance care quality—a critical goal for achieving a more efficient and effective healthcare system.

## Data Availability

All data produced in the present study are available upon reasonable request to the author

